# Sedation practices in patients intubated in the emergency department compared to the intensive care unit

**DOI:** 10.1101/2024.03.26.24304926

**Authors:** Jariya Sereeyotin, Christopher Yarnell, Sangeeta Mehta

**Affiliations:** Department of Anesthesiology, Division of Critical Care Medicine, King Chulalongkorn Memorial Hospital and Faculty of Medicine, Chulalongkorn University, Bangkok, Thailand; Department of Medicine, Sinai Health; Interdepartmental Division of Critical Care Medicine, University of Toronto, Toronto, ON, Canada; Department of Critical Care Medicine and Research Institute, Scarborough Health Network, Toronto, ON, Canada

## Abstract

**Purpose:** This study aimed to compare sedation management during and after intubation in the emergency department (ED) versus the intensive care unit (ICU).

**Methods:** This was a single-center retrospective cohort study of adults intubated in the ED or in the ICU and received mechanical ventilation between January 2018 and February 2022. We collected data from the electronic medical record. The primary outcome was duration from intubation to first documentation of light sedation, defined as a Sedation Agitation Scale score (SAS) of 3-4.

**Results:** The study included 264 patients, with 95 (36%) intubated in the ED and 169 (64%) in the ICU. Regarding anesthetic agents used for intubation, ketamine was the most commonly used drug in the ED and was used more frequently than in the ICU (61% vs 40%, p=0.001). Propofol was the predominant sedative used in the ICU, with a higher prevalence compared to the ED (50% vs 33%, p=0.01). Additionally, benzodiazepines and fentanyl were more frequently used in the ICU (39% vs 6%, p<0.001 and 68% vs 9.5%, p<0.001, respectively). Within 24 hours after intubation, 68% (65/95) ED patients and 82% (138/169) patients intubated in ICU achieved light sedation, with median durations of 13.5 hours and 10.5 hours. Patient location in the ED at intubation was associated with decreased probability of achieving light sedation at 24 hours (adjusted odds ratio 0.64, p=0.04).

**Conclusion:** Critically ill patients intubated in the ED are at risk of deeper sedation and a longer time to achieve light sedation compared to patients intubated in the ICU.

## Introduction

Effective sedation management is crucial for facilitating intubation, ensuring comfort, and promoting patient-ventilator synchrony in mechanically ventilated adults. However, this management poses challenges in both the emergency department (ED) and the intensive care unit (ICU) due to limited physiological reserves and the potential for serious complications in critically ill patients.[1, 2] Moreover, inappropriate or excessive sedation has been associated with adverse outcomes in critically ill patients.

Several studies have reported that deep sedation in mechanically ventilated patients during the first 48 hours after ICU admission is associated with a higher risk of death and delirium, as well as delayed time to extubation.[3–7] Despite the 2018 Pain, Agitation/Sedation, Delirium, Immobility and Sleep Disruption (PADIS) guidelines[8] recommending light sedation in critically ill mechanically ventilated adults, deep sedation is commonly used for intubated patients in the ED. In a recent multicenter, prospective cohort study of 324 patients receiving mechanical ventilation in the ED, 52.8% of intubated patients were deeply sedated [defined as Sedation Agitation Scale (SAS) score of 1 or 2], and deep sedation continued throughout the first 48 hours of ICU admission.[9] Therefore, an early emphasis on sedation minimization in critically ill patients may be beneficial.

Many critically ill patients are initially intubated and managed in the ED, yet there are few studies evaluating sedation practices for mechanically ventilated patients in the ED.[9, 10] The overall purpose of our study was to compare sedation management during and after intubation in patients intubated in the ED versus those intubated in the ICU. Given that our study period included the COVID-19 pandemic, during which rapid sequence intubation (RSI) was recommended for all patients with suspected or confirmed COVID-19 infection[11, 12], we also explored the change in sedation management prior to and during the COVID-19 pandemic. Moreover, we aimed to identify patient factors and other potentially modifiable factors associated with deep sedation to improve sedation management in critically ill patients.

## Materials and Methods

This retrospective cohort study was conducted at a tertiary care university-affiliated hospital and approved by the Sinai Health Research Ethics Board (REB#23-0002-C) on February 15^th^, 2023. Due to the retrospective research design and the fact that no identifying information would be collected, the need for informed consent was waived. We identified eligible consecutive patients from an ICU research screening database and electronic medical record (EMR) prior to (January 1^st^, 2018-January 31^st^, 2020) and during the COVID-19 pandemic (February 1^st^, 2020-February 28^th^, 2022). We accessed individual patient information between February 15^th^, 2023 and August 31^st^, 2023; however, the identifying information was not recorded during data collection.

The study included adult patients aged 18 years or older who were intubated either in the ED or within the first 24 hours of ICU admission and received mechanical ventilation. Exclusion criteria were: need for deep sedation or neuromuscular blocking agents after intubation (e.g., therapeutic hypothermia after cardiac arrest, moderate to severe acute respiratory distress syndrome, or status epilepticus); death within 48 hours of intubation; transferred to another department after intubation; and not expected to achieve light sedation within 48 hours (e.g., intracranial hemorrhage, brain stem infarction, or encephalopathy).

The primary outcome was the time from intubation to the first documentation of light sedation, defined as SAS 3-4. Secondary outcomes included SAS at 6, 12, 24 and 48 hours after intubation; intubation technique (RSI and non-RSI; RSI was defined as the administration of sedative agents followed by rapid onset neuromuscular blocking agents without positive pressure ventilation unless it was necessary); drug types and dosage for intubation and the postintubation period; potential factors associated with deep sedation after intubation; and incidence of hemodynamic instability (defined as hypotension SBP < 90 mmHg or at least 20% decrease of initial MAP for > 30 minutes; new requirement for or increase in dose of vasopressors; fluid bolus > 15 ml/kg to maintain target blood pressure; or new onset of arrhythmia) within 30 minutes from the start of intubation.

### Statistical analysis

Demographic variables were presented by descriptive analysis, using frequency (percentage) for categorical data. For continuous data, Kolmogorov-Smirnov was used for normality testing and data expressed as mean (standard deviation, SD) or median (interquartile range, IQR) as appropriate. The primary outcome was analyzed using a Cox proportional hazards model that adjusted for relevant covariates. Covariates included age, Acute Physiologic Assessment and Chronic Health Evaluation (APACHE) II score, obesity (body mass index ≥ 30 kg/m^2^), renal insufficiency (defined as GFR < 30 mL/min/1.73m^2^), end-stage liver disease, baseline Glasgow Coma Scale (GCS) and potential confounders for achieving light sedation including administration of benzodiazepines and neuromuscular blocking agents after intubation. From the primary analysis we reported the odds ratio of achieving light sedation according to patient location at the time of intubation (ED versus ICU).

For the secondary analysis, we used logistic regression models to identify potential predictors for the following binary outcomes: RSI versus non-RSI, deep sedation versus light sedation, presence of hemodynamic instability versus none.

All analyses were performed using SPSS software version 28 and a two-sided P value < 0.05 was considered statistically significant.

## Results

Between January 2018 and February 2022, a total of 314 patients were eligible for the inclusion criteria, of whom 50 patients were excluded due to the reasons as shown in Fig 1. Ultimately, 264 patients were included in the primary analysis, with 95 (36%) intubated in the ED and 169 (64%) intubated in the ICU. The mean (SD) age was 60 (18) years and 153 (58%) were male. The two groups were similar at baseline prior to intubation, other than a higher proportion of patients with GCS ≤ 8 in the ED group (68% vs 25%), and higher mean age in the ICU group (63 vs 56 years). The main reason for intubation in the ED was neurological dysfunction while respiratory failure was the most common reason for intubation in the ICU. Patient characteristics are shown in Table 1.

**Fig 1.**
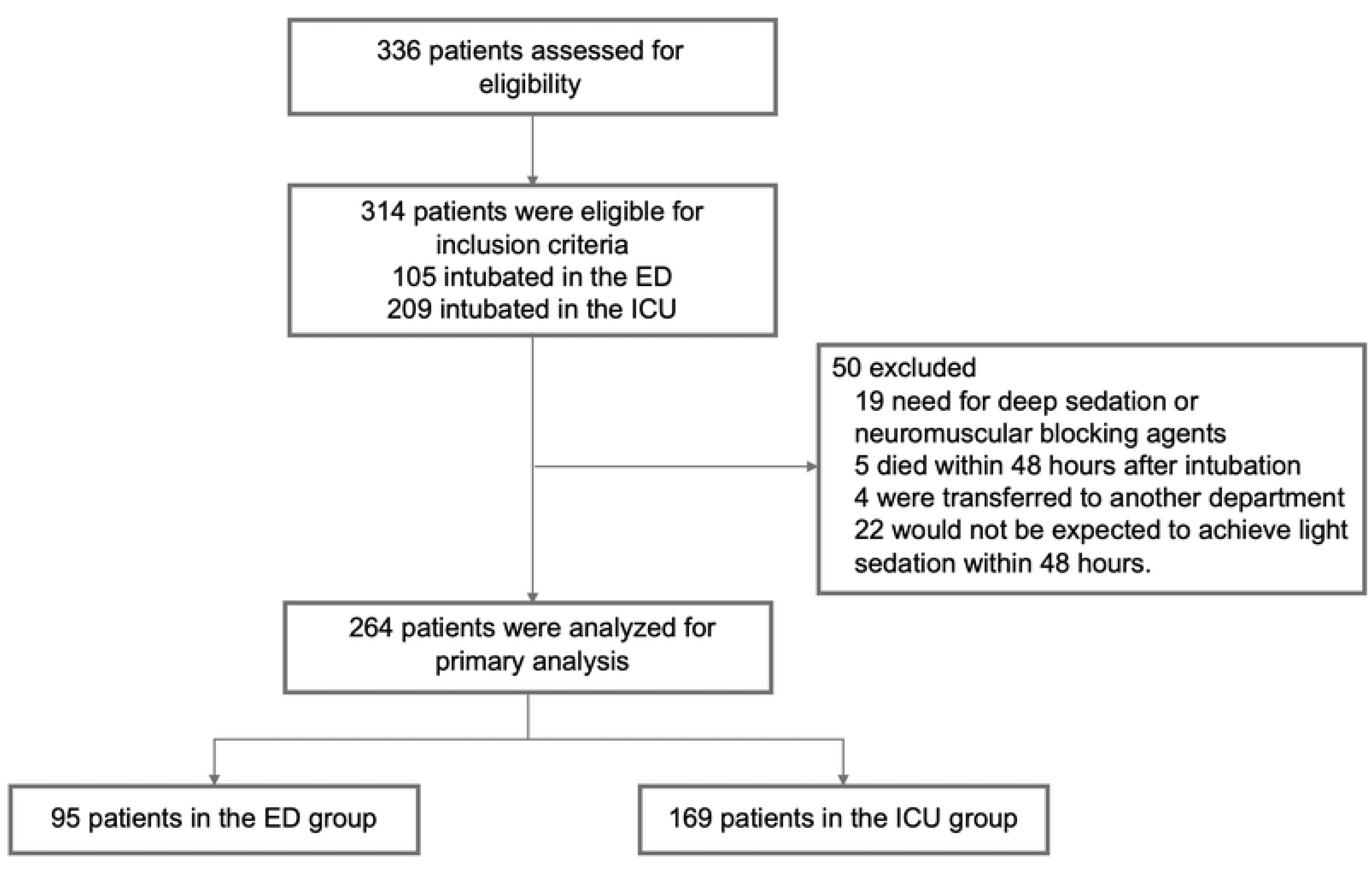
Flow of participants in a study of the sedation practices in patients intubated in the emergency department compared to the intensive care unit. ED = Emergency Department, ICU = Intensive Care Unit.

**Table 1.** Patient characteristics prior to endotracheal intubation.

| **Variables** | **Emergency**<br>**Department N=95** | **Intensive Care**<br>**Unit N=169** | **P value** |
| --- | --- | --- | --- |
| Age, years, mean (SD) | 56 (21) | 63 (15) | 0.004 |
| Male sex, n (%) | 55 (58) | 98 (58) | 0.99 |
| BMI (kg/m<sup>2</sup>), median (IQR) | 26 (21-28) | 26 (23-31) | 0.30 |
| Race, n (%)* |  |  |  |
| White | 68 (72) | 110 (65) | 0.34 |
| Black | 9 (9) | 10 (6) |  |
| Other person of color | 18 (19) | 49 (29) |  |
| APACHE II, median (IQR) | 22 (17-27) | 22 (17-27) | 0.80 |
| Baseline GCS before intubation |  |  |  |
| GCS ≤ 8, n (%) | 61 (68) | 36 (25) | <0.001 |
| GCS > 8, n (%) | 29 (32) | 110 (75) |  |
| COVID infection, n (%) |  |  |  |
| Positive | 6 (6) | 6 (4) | 0.30 |
| Negative | 89 (94) | 163 (96) |  |
| Underlying disease, n (%) |  |  |  |
| Hypertension | 34 (36) | 72 (43) | 0.28 |
| Diabetes mellitus | 26 (27) | 40 (24) | 0.51 |
| COPD | 9 (9) | 20 (12) | 0.56 |
| Renal insufficiency | 12 (13) | 28 (17) | 0.39 |
| Hepatic dysfunction | 10 (11) | 27 (16) | 0.22 |
| End-stage liver disease | 0 (0) | 3 (2) | 0.19 |

| Reason for intubation, n (%) |  |  |  |
| --- | --- | --- | --- |
| Airway obstruction | 8 (8) | 11 (7) | 0.56 |
| Respiratory failure | 25 (26) | 107 (63) | <0.001 |
| Cardiogenic shock | 4 (4) | 4 (2) | 0.40 |
| Neurological dysfunction | 46 (48) | 22 (13) | <0.001 |
| Sepsis | 3 (3) | 24 (14) | 0.004 |
| Cardiac arrest | 7 (7) | 4 (2) | 0.05 |
| Other | 2 (2) | 0 (0) | 0.06 |

\* Race was visually identified and recorded in the ICU research database by the research coordinator.
BMI = Body Mass Index; APACHE II = Acute Physiologic Assessment and Chronic Health Evaluation II score; GCS = Glasgow Coma Scale; COPD = Chronic Obstructive Pulmonary Disease; SD = standard deviation; IQR = interquartile range

### Primary outcome and sedation agitation scale

Within the first 24 hours after intubation, 68.4% (65/95) ED patients and 81.7% (138/169) ICU patients achieved light sedation, with median durations of 13.5 hours and 10.5 hours, respectively. Patient location in the ED at the time of intubation was associated with decreased probability of achieving light sedation at 24 hours, with adjusted odds ratio of 0.64 (p=0.04; 95% CI, 0.42 to 0.97) (S1 Table in S1 Appendix). Cumulative hazard curves assessing time from intubation to the first documentation of achieving light sedation are shown in Fig 2.

**Fig 2.**
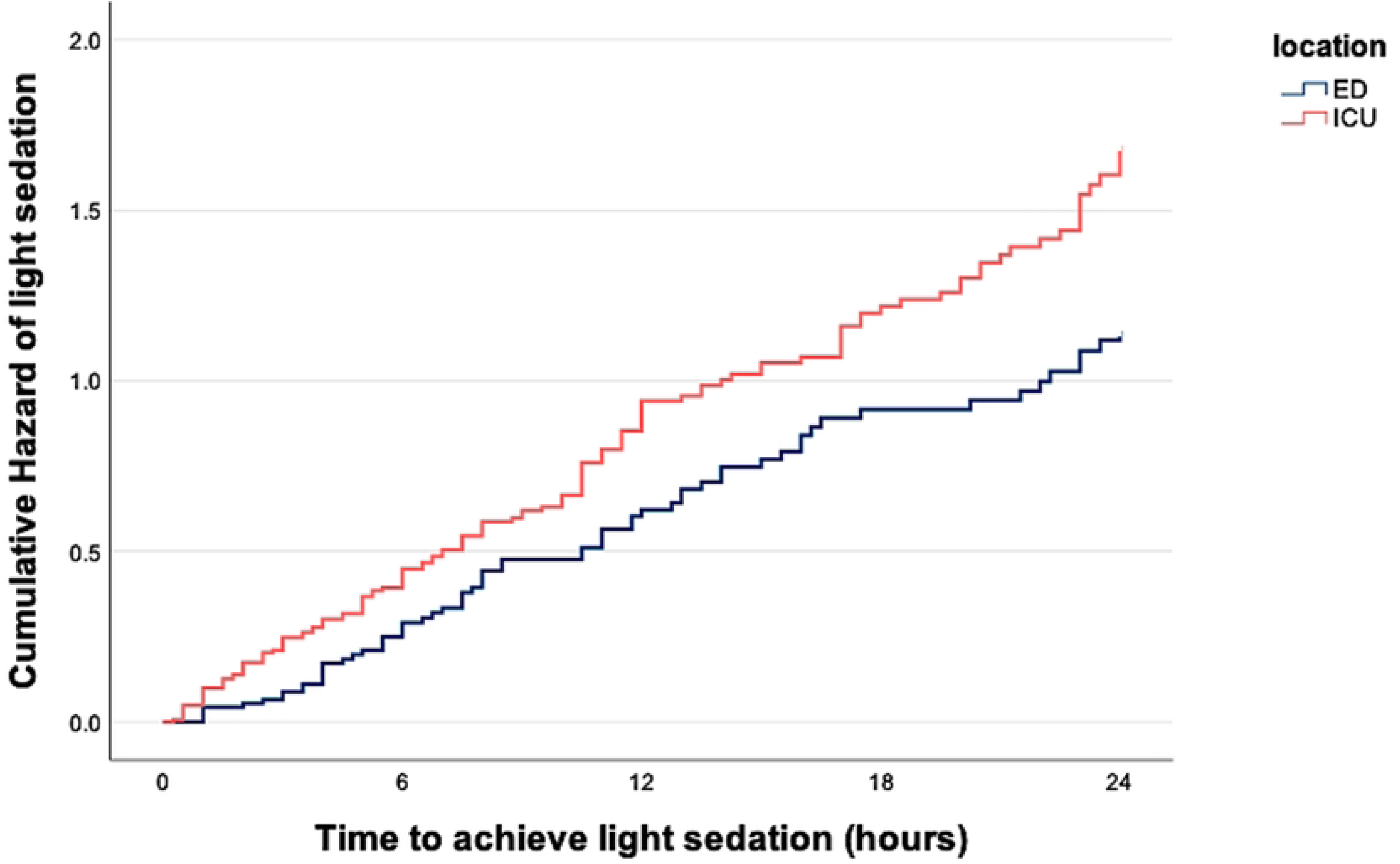
Cumulative hazard of the time to achieve light sedation at 24 hours, by patients’ location of intubation. Within the first 24 hours after intubation, the median time to achieve light sedation was 13.5 hours for patients intubated in the ED and 10.5 hours for those intubated in the ICU. The mean odds ratio for achieving light sedation at 24 hours was 0.69 for the ED (p=0.01; 95% confidence interval [CI], 0.51-0.92). Deep sedation (SAS 1 or 2) was more frequently observed in the ED group at all time points, especially at 12 and 48 hours (60.2% vs. 46.3%, p =0.03; and 26.5% vs. 13.0%, p=0.02, respectively) (Fig 3). More detail for the number of patients who were deeply sedated and lightly sedated at each time point is reported in S2 Table in S1 Appendix.

**Fig 3.**
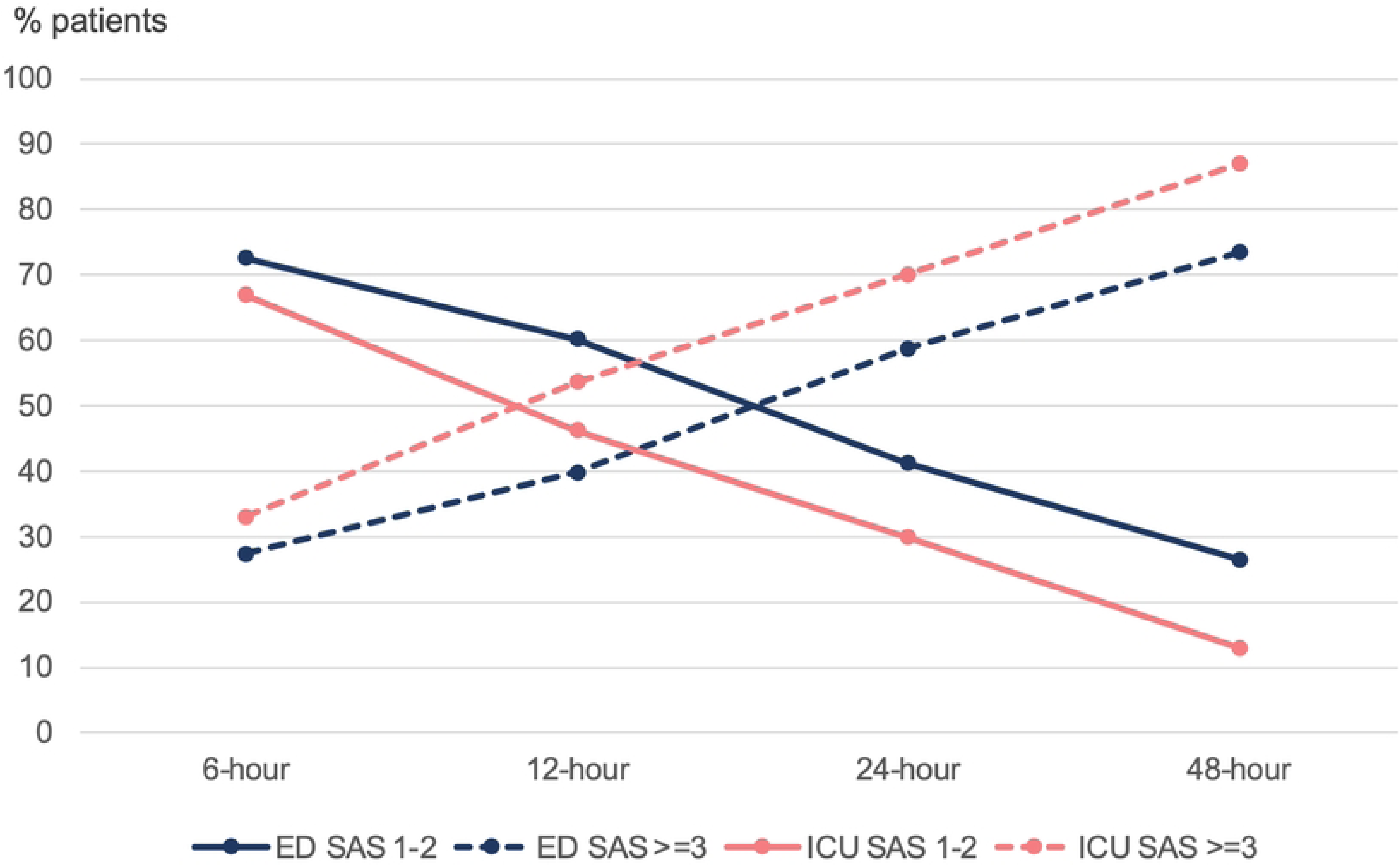
Sedation Agitation Scale at each time point in ED and ICU patients. The Sedation Agitation Scale (SAS) at each time point is shown for patients intubated in the ED and the ICU. Deep sedation (SAS 1 or 2) was more frequently observed in the ED group, and a higher prevalence of light sedation (SAS 3 or 4) was found in the ICU group at all time points, with statistically significant differences at 12 and 48 hours (60.2% vs. 46.3%, p=0.03; and 26.5% vs. 13.0%, p=0.02, respectively for deep sedation; 39.8% vs. 53.7%, p =0.03 and 73.5% vs. 87.0%, p=0.02, respectively for light sedation).

### Intubation technique, drug types and drug dosages

RSI was significantly more frequent in the ED compared to the ICU (82.1% vs. 30.2%, p <0.001). During the COVID-19 pandemic, the incidence rate of RSI was significantly increasing from 75% to 92.3% (p=0.03) in the ED and from 17.3% to 50.8% in the ICU (p<0.001). Intubation technique prior to and during the COVID-19 pandemic in the ED and ICU appear in S3 Table and S1 Fig in S1 Appendix.

Regarding anesthetic agents used for intubation (Table 2), ketamine was the most commonly used drug in the ED and was used more frequently than in the ICU (61% vs 40% patients, p=0.001). Propofol was the predominant sedative used in the ICU, with a higher prevalence compared to the ED (50% vs 33%, p=0.01). Additionally, benzodiazepines, opiates and topical xylocaine were more frequently used in the ICU (39% vs 6%, p 0.001; 68% vs 10%, p<0.001; and 15% vs 3%, p=0.003; respectively).

**Table 2.**
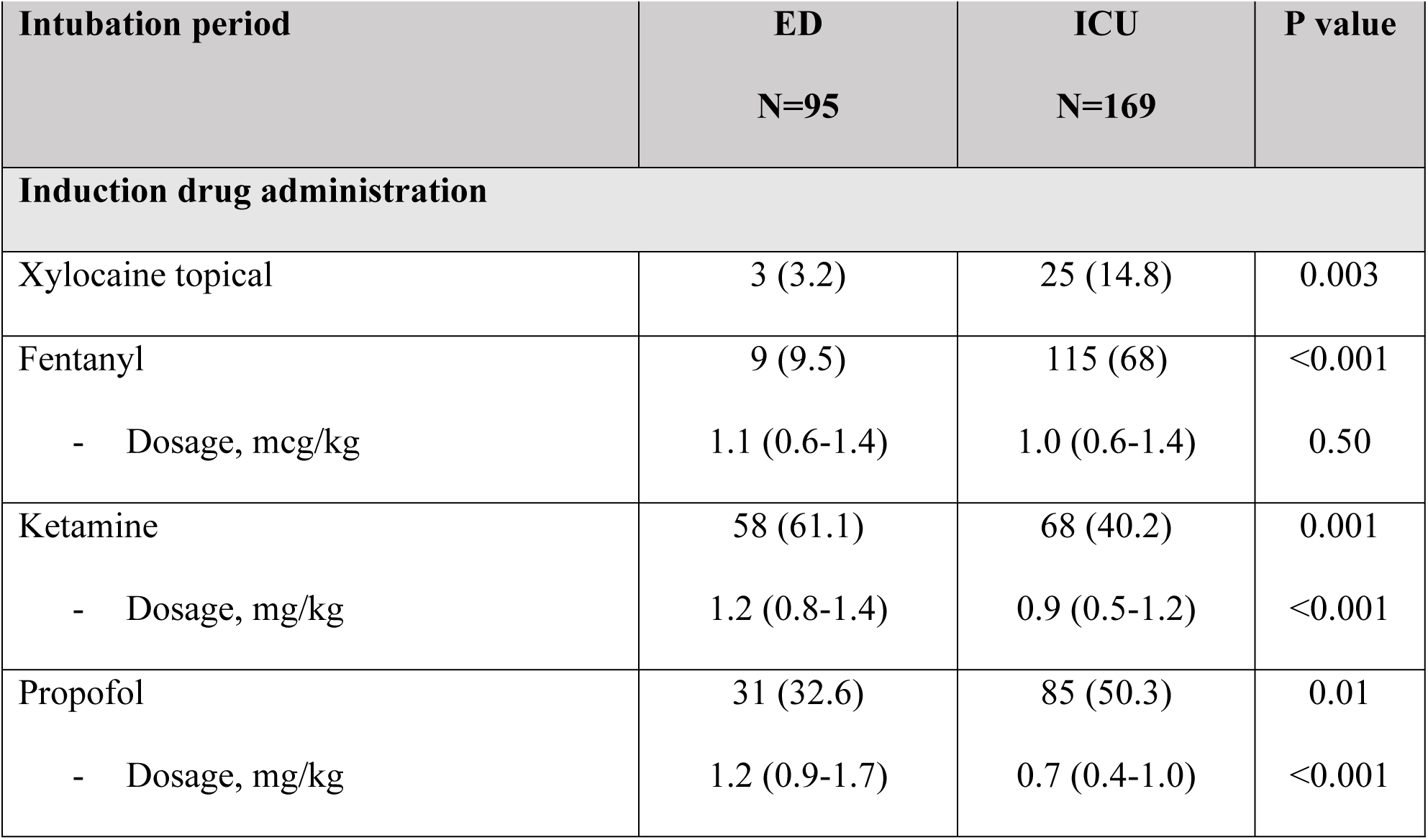
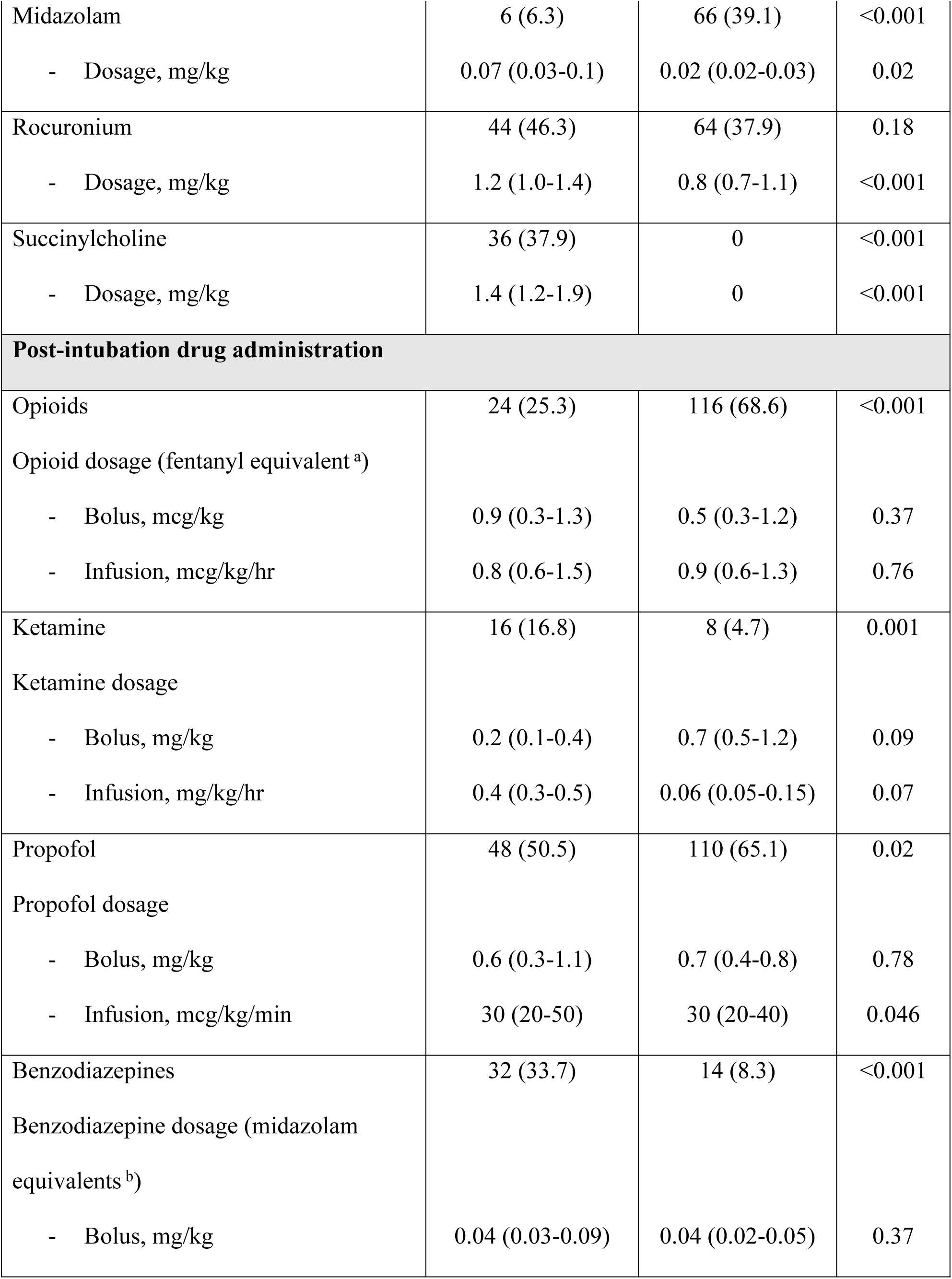
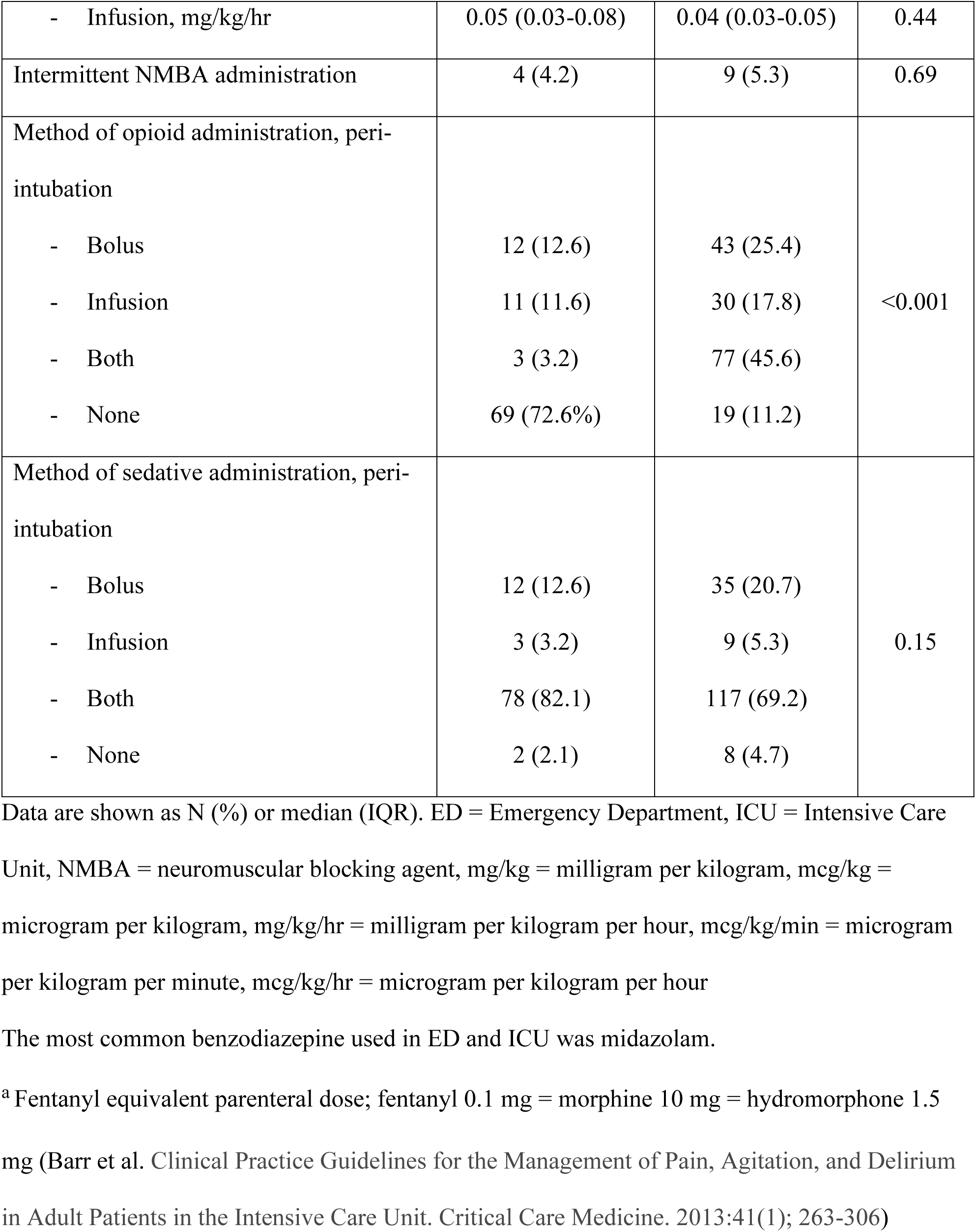
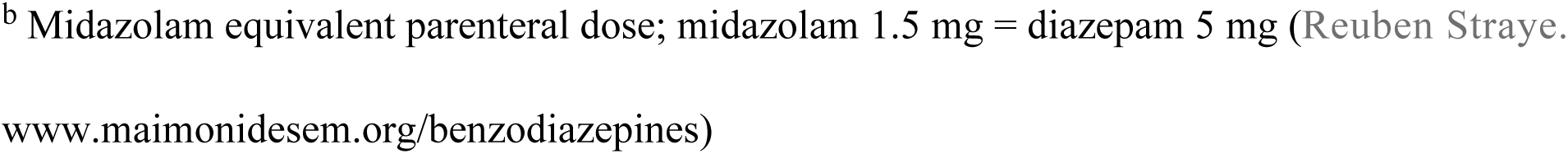
Sedative, analgesic, and paralytic medications used for intubation and post-intubation in the ED versus ICU.

In terms of drug dosages, higher doses of sedatives and neuromuscular blocking agents were used for intubation in the ED, while no significant difference in opioid dosage was found. We identified 3 patients intubated in the ED who received neuromuscular blocking agents alone without documented sedatives: two were diagnosed with drug intoxication and had baseline GCS 3 with no improvement after therapeutic naloxone administration, and the other had baseline GCS 5 and was intubated due to COVID-19 infection and septic shock.

After intubation, opioids were less commonly used (25.3% vs. 68.6%, p <0.001), while ketamine and benzodiazepines were more frequently used (16.8% vs 4.7%, p=0.001; and 33.7% vs. 8.3%, p <0.001, respectively) in the ED compared to the ICU. The bolus and infusion dosages were similar in ED and ICU except for a higher dose of propofol infusion in the ED [30 (20-50) vs 30 (20-40) mcg/kg/min, p=0.046].

Compared to prior to COVID-19 pandemic, rocuronium was used for intubation at higher doses both in the ED and the ICU during the COVID-19 pandemic [0.9 (0.6-1.0) vs 1.3 (1.2-1.6) mg/kg, p<0.001 for ED; and 0.7 (0.6-0.8) vs 1.1 (0.7-1.4) mg/kg, p<0.001 for ICU]. After intubation, higher prevalence of using propofol and benzodiazepines as well as higher doses of propofol infusion were observed in the ICU while higher prevalence of ketamine was observed in the ED (S4.1 and S4.2 Tables in S1 Appendix).

Following intubation in the ICU, SAS scores were documented in the patient record every 4 hours. In the ED, GCS was recorded before transfer to the ICU, whereas no sedation assessment tools were documented.

### Other outcomes

A multivariable logistic regression analysis was performed to identify the effects of APACHE II score, baseline GCS, renal insufficiency, end-stage liver disease, use of midazolam for induction and sedative administration methods (bolus, infusion, or both) on the likelihood that patients would be deeply sedated 24 hours after intubation. In adjusted analysis, patients with renal insufficiency (GFR < 30 mL/min/1.73m^2^) were 2.78 times (p=0.03) more likely to be deeply sedated at 24 hours than those without renal insufficiency. In addition, patients with baseline GCS ≤ 8 were 2.17 times (p=0.03) more likely to be deeply sedated than those with baseline GCS > 8 (S5 Table in S1 Appendix). Neurologic dysfunction as a reason for intubation and being intubated during the COVID-19 pandemic were two independent factors strongly associated with greater use of the RSI technique (OR 2.9, p<0.001; and OR 3.6, p<0.001, respectively) (S6 Table in S1 Appendix).

Regarding outcomes after intubation, the incidence of cardiovascular instability requiring vasopressor treatment was higher in the ICU group compared to the ED group (68.6% vs. 45.3%, p<0.001) (Table 3). Increasing age, a diagnosis of sepsis and fentanyl use for induction were found to be associated with the risk of cardiovascular instability, while sedative agents used for induction, including propofol, ketamine, and benzodiazepine, were not (S7 Table in S1 Appendix). Days of mechanical ventilation, hospital length of stay and 28-day mortality were higher in the ICU group compared with the ED group (4 (2-9) vs 2 (1-3) days, p<0.001; 14 (8-47) vs 8 (3-16) days, p<0.001 and 37.7% vs. 12.3%, p<0.001, respectively). However, patients who were deeply sedated at any time point had longer days of mechanical ventilation, longer hospital length of stay and fewer ventilator-free days at 28 days than the lightly sedated group (Fig 4). In addition, deeply sedated patients were less likely to be discharged home and were more commonly transferred to a rehabilitation facility than the lightly sedated group. More details are provided in S8 Table in S1 Appendix

**Fig 4.**
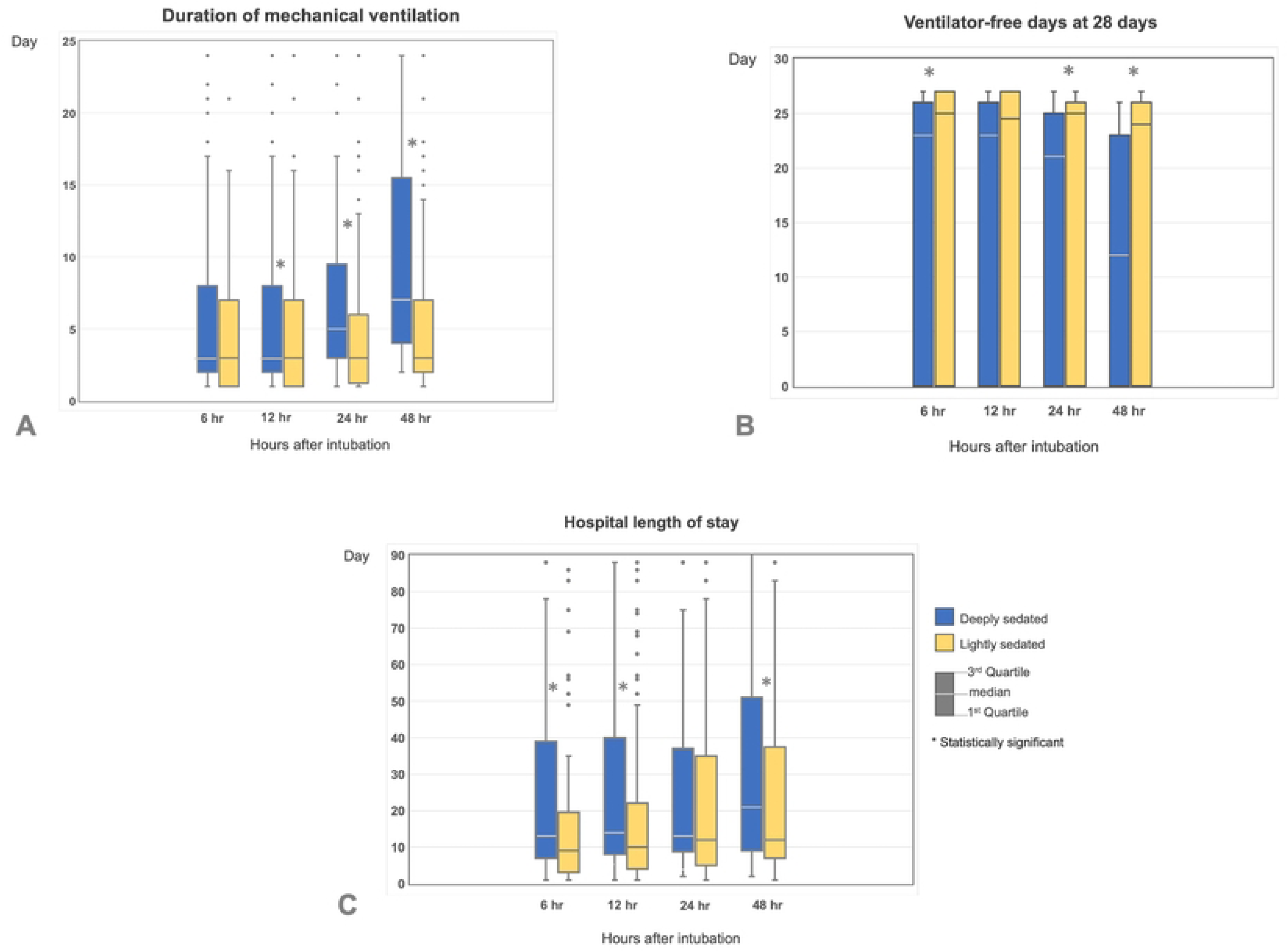
Duration of mechanical ventilation, ventilator-free days at 28 days, and hospital length of stay between deeply sedated and lightly sedated groups. Deeply sedated is defined as SAS 1-2; Lightly sedated is defined as SAS ≥3., * statistical significance A. Duration of mechanical ventilation was longer in deeply sedated group compared to lightly sedated group at any time point with statistical significance at 12, 24 and 48 hours after intubation., B. Patients who were lightly sedated at 6, 24 and 48 hours after intubation had more ventilator-free days at 28 days than those who were deeply sedated., C. At 6, 12 and 24 hours after intubation, deeply sedated patients had significantly longer hospital length of stay than lightly sedated patients.

**Table 3.**
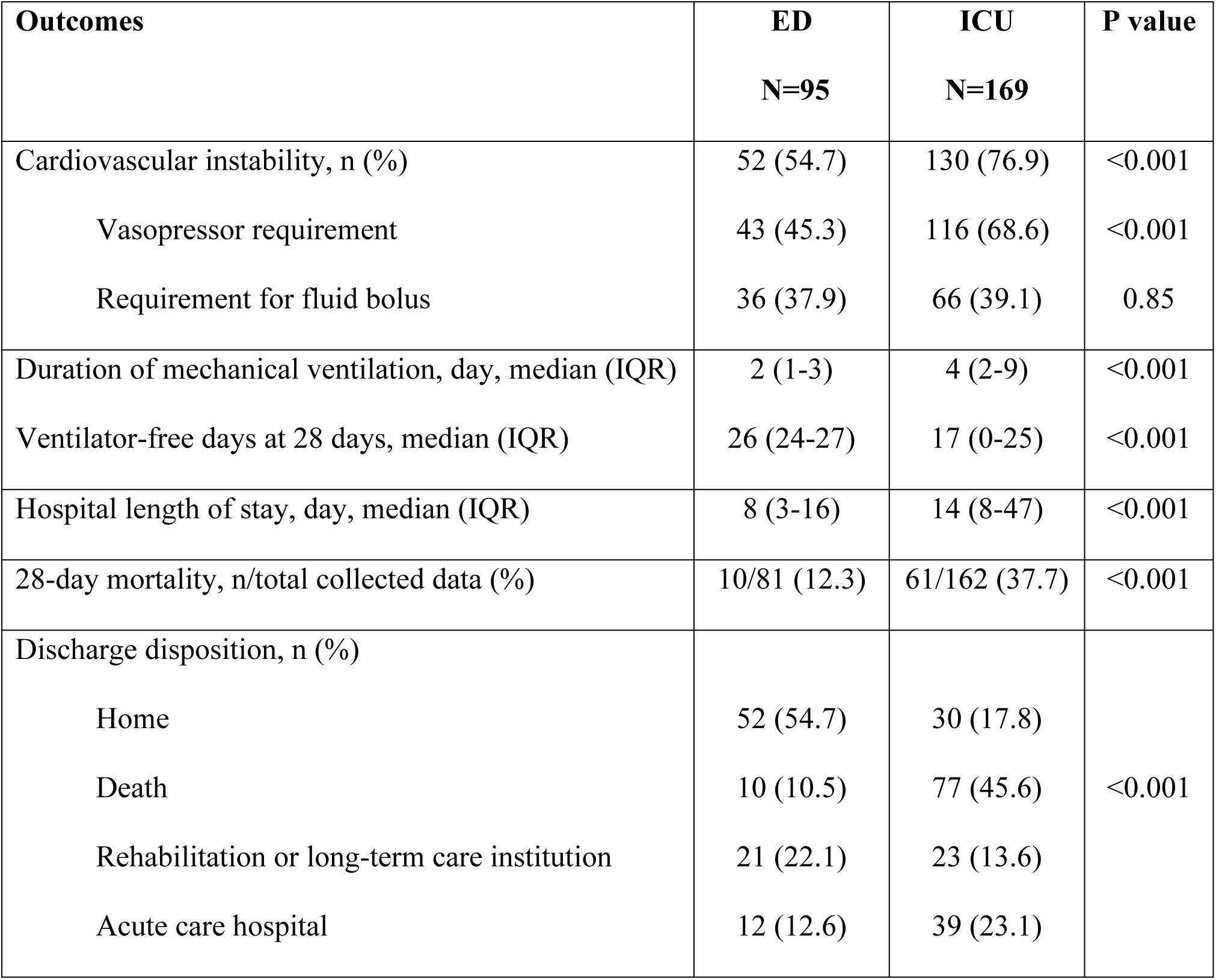
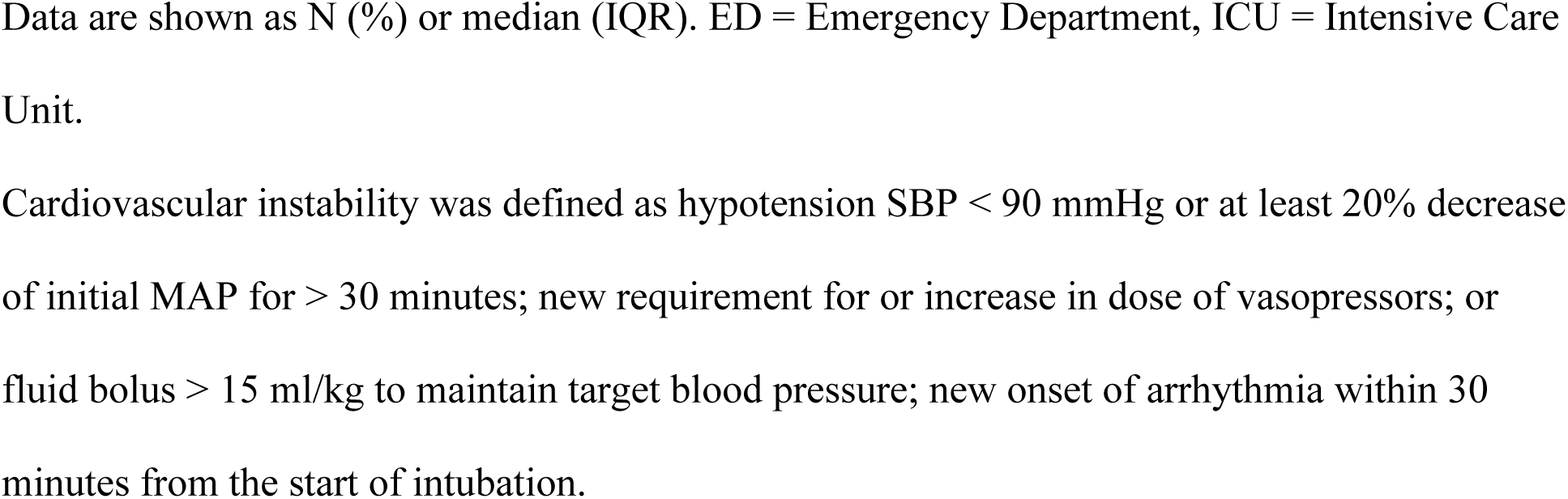
Cardiovascular instability and ICU outcomes between the ED and ICU group.

## Discussion

In this single-center retrospective cohort study, patients who were intubated and had initiation of invasive ventilation in the ED had a longer time to achieve light sedation than patients intubated in the ICU. Moreover, deep sedation was more frequently observed in the ED group during the first 48 hours of mechanical ventilation.

Our study emphasizes the impact of ED sedation practice for mechanically ventilated patients on ICU outcomes, including fewer ventilator-, ICU- and hospital-free days.[9, 10] Our results are consistent with the ED-SED study, which evaluated ED sedation practices in mechanically ventilated patients.[9] They found that deep sedation was frequently started in the ED and continued throughout the first 48 hours of ICU admission. More than 70% of deeply sedated patients in their ICU received sedation management starting from the ED. Likewise, this study also found a higher frequency of deep sedation in patients intubated in the ED at 6, 12, 24 and until 48 hours after intubation compared to those intubated in the ICU.

There is limited high-quality evidence to clarify the optimal dose of sedatives for intubation in critically ill adults.[13–15] Higher doses of sedatives and neuromuscular blocking agents were used for intubation in the ED despite the finding that the most common indication for intubation in the ED was neurologic dysfunction (48%). Concerningly, 3 patients with low baseline GCS (3-5) were intubated in the ED with the administration of neuromuscular blocking agents alone without sedation, highlighting an opportunity for improvement.

In a study of intubation practice in 3,659 critically ill patients across 29 countries, the main reasons for intubation were respiratory failure (52%) and neurological dysfunction (31%), similar to our study (50% respiratory failure, 26% neurological dysfunction). In their study the most common drugs used for induction were propofol (41.5%), midazolam (36.4%), etomidate (17.8%) and ketamine (14.2%). Propofol was significantly associated with cardiovascular instability compared with etomidate (64% vs 50% respectively, absolute difference 14%; 95% CI 1.4-27.0%, p=0.02) when administered to patients with hemodynamic instability.[15] In our study, ketamine was the most commonly used drug in the ED (61.1%), while propofol and ketamine were the two most predominant sedatives used in the ICU (50.3% and 40.2%, respectively). The use of specific sedatives for induction was not found to be a risk factor for deep sedation at 24 hours nor for cardiovascular instability.

Regarding postintubation sedation, 33.7% of patients in the ED received benzodiazepines, compared with 8.3% in the ICU. In addition, only 25.3% of patients in the ED group received opioids for postintubation analgesia. These sedation practices were inconsistent with the 2018 PADIS Guidelines[8], which recommend an assessment-driven, protocol-based, stepwise approach for pain and sedation management in critically ill adults, using analgesia first. Moreover, benzodiazepines are not recommended for sedation.[16, 17] Another area for improvement is that sedation assessment tools should be used in the ED to better target light sedation.

Our logistic regression analysis identified that patients with renal insufficiency and patients with low baseline GCS (≤8) were more likely to have deep sedation following intubation,[18–20] suggesting that a cautious approach to sedative administration may be wise in these groups. Furthermore, it was not surprising that the COVID-19 pandemic and neurologic dysfunction as the reason for intubation were independent factors strongly associated with performing RSI technique. The most likely reason for patients with neurological dysfunction was concern for increased intracranial pressure during laryngoscopy.

Regarding outcomes after intubation, increasing age, sepsis diagnosis and pretreatment with fentanyl were associated with cardiovascular instability in our study, which is consistent with prior studies.[21–23] This could explain the higher incidence of cardiovascular instability in the ICU group. Although this study has not shown an association between deep sedation and mortality outcome as previous studies did,[5, 6, 10, 24] we found that fewer patients were discharged home and more were discharged to a rehabilitation hospital in the early deep sedation group. Moreover, fewer ventilator-free days and longer hospital length of stay were observed in this group.

To our knowledge, this is the first study comparing sedation practices for intubation and post-intubation between the ED and the ICU. Strengths of this study include detailed data on drug types and dosages, evaluation during and prior to the COVID-19 pandemic, complete patient follow-up, and identification of predictors of deep sedation. This study has limitations. First, it was a single-center study, so these data may not represent broader clinical practice. However, the prevalence of deep sedation in the ED in our study was similar to previous studies. Second, the duration from intubation to the first documentation of light sedation was analyzed for the primary outcome, which mainly depended on the frequency of nurse assessments and might not be the actual time for achieving light sedation. For this reason, we also collected the sedation score at four specific time points. The data was in the same direction: a higher frequency of deep sedation at every time point and a longer time required to achieve light sedation were found in the ED group. Third, unmeasured confounders between the location of intubation and the use of post-intubation sedation could partially explain our findings. Finally, due to the retrospective study design, the association between deep sedation and longer-term outcomes (e.g., 90-day mortality, cognitive outcomes, etc.) could not be evaluated.

## Conclusion

Patients intubated in the ED were more deeply sedated and took longer to achieve light sedation than patients intubated in the ICU.

## Data Availability

All relevant data are within the manuscript and its Supporting Information files.

## Acknowledgements

We thank Sumesh Shah, research coordinator and Stanley Oei, respiratory therapist for sharing their screening database.

## Supporting information

**S1 Appendix** Table E1-E8 and Fig E1

